# Beyond Mortality: The Importance of Morbidity Data for Understanding Pandemics - The Case of the 1918–1920 Influenza Pandemic in Switzerland

**DOI:** 10.64898/2025.12.15.25342287

**Authors:** Katarina L Matthes, Sofie Jörg, Rick J Mourits

## Abstract

Historical morbidity data from past epidemics and pandemics are often sparsely or not available at the individual level. As a result, most studies rely on mortality data to assess the dynamics and severity of epidemics or to examine demographic and social disparities. Although mortality inarguably reflects the severity of an epidemic, it only tells half the story, and a comprehensive understanding requires consideration of both morbidity and mortality. This paper harmonizes morbidity data across different administrative levels and demonstrates how spatial analysis can be used to examine demographic and socioeconomic differences between morbidity and mortality. Linking aggregated data to explanatory factors at the ecological level and applying spatial regression analysis enables the investigation of disparities between morbidity and mortality even in the absence of individual-level data. The findings indicate that the factors shaping morbidity patterns differ from those influencing mortality, underscoring the importance of jointly examining both dimensions to fully understand pandemic dynamics. Hence, this paper encourages historical demographers and historical epidemiologists to consider studying morbidity data rather than dismissing it due to perceived limitations in data quality.

## 1. Introduction

Historical morbidity data from past epidemics and pandemics are often limited or unavailable at the individual level. Therefore, most studies rely on mortality data to assess the dynamics and severity of epidemics or pandemics, as well as to examine demographic and social disparities. Mortality data are more frequently preserved and, in many cases, are available at the individual level through death records. In contrast, morbidity data present several challenges. Individual-level information is often missing due to the lack of comprehensive health registers, and if available data are typically aggregated across different administrative levels. Moreover, during peak periods of a pandemic, reporting at lower administrative levels may break down, with data often being consolidated and reported directly at higher levels. This reduces the granularity and accuracy of the data, making detailed analysis more difficult.

Morbidity data nonetheless remains essential to any comprehensive understanding of pandemics. While mortality captures the most extreme dimension of a pandemic’s severity, it does not fully reflect the burden of disease, a complete assessment demands attention to both. Focusing solely on death rates obscures how many people are affected by illness and the long-term consequences they may carry, overlooking important demographic and social disparities in how populations experience sickness versus death. For example, older adults may face prolonged disease even if deaths are concentrated elsewhere, socially disadvantaged groups can carry a heavier disease burden that mortality alone does not capture or women often experience higher levels of illness than men despite living longer, also called as ‘male-female health-survival paradox’ ^1^. While the ‘male-female health-survival paradox’ has come more into focus in recent decades ^2,3^, there is little evidence to support it based on historical data. Without morbidity data, these patterns remain hidden, leaving an incomplete picture of a pandemic’s true characteristics and possible effects. For the 1918-1920 pandemic, a few local studies have shown indications of differences between mortality and morbidity in demographic groups. For example, a survey of the 1918–1919 pandemic in the United States showed that during the first wave, children aged 0–9 were disproportionately affected, even though they did not experience the highest mortality ^4^. Similarly, morbidity in influenza pandemics was reported to be higher among women, while mortality was higher among men ^4^. Most studies of the past influenza pandemics were based on mortality data, which often led to the assumption that men faced a higher risk of disease ^5,6^. However, evidence suggests that women were more affected by the pandemic in terms of illness, even if not in terms of death ^7,8^, a pattern also observed in a 1919 medical survey of Swiss factory employees, indicating a higher morbidity rate among female workers compared to male workers ^9^.

Taken together, morbidity data are a crucial contribution to a comprehensive understanding of pandemics, yet they are often harder to obtain and analyse than mortality data. When examining differences and their determinants beyond the local level, spatial data can prove particularly useful. Aggregated spatial morbidity data can be linked to ecological factors such as population density by age and sex, socioeconomic indicators, or access to healthcare, and compared with corresponding mortality data. Although such analyses do not operate at the individual level, they can still offer a valuable comprehensive overview of a pandemic’s morbidity and mortality.

In the past, some scholars have argued against using morbidity data, particularly from the time of the 1918-1920 pandemic, because it was underreported and considered unreliable^10,11^ . While this may be true when trying to determine total infection numbers, a challenge that also applies to more recent pandemics, morbidity data can still provide valuable insights when analysed at the spatial level. By comparing morbidity patterns across regions and relating them to mortality data, as well as to assumed disease trajectories, these data can offer a meaningful understanding of the data quality as well as the pandemic’s development and impact, even if absolute case counts are uncertain. Ecological determinants can be linked to the spatial morbidity and mortality data, enabling an examination of disparities between the two, even where individual-level data are unavailable.

When analysing data at fine geographic scales, we can utilise that morbidity and mortality are rarely distributed randomly across space. Morbidity and mortality often exhibit spatial autocorrelation, meaning nearby areas tend to share similar patterns due to common environmental or social conditions, a structure that standard linear regression models may fail to capture. By situating morbidity and mortality within their geographic context, spatial analysis enhances analytical robustness and deepens our understanding of historical morbidity and mortality patterns. However, spatial heterogeneity, or non-stationarity, can complicate matters as the factors driving clustering in one region may differ substantially from those operating elsewhere. Ignoring these spatial structures risks producing biased estimates and misleading conclusions, since traditional statistical methods assume independence and homogeneity across space. For these reasons, methods of spatial analysis are essential to estimate the relation of spatial morbidity or mortality and determinates.

There are only few contemporary studies examining morbidity^12–14^, and to our knowledge, none have analysed the spatial distribution and dynamics of morbidity data, as most research focuses solely on mortality. In this paper, we present a method for harmonizing morbidity data across different administrative levels and demonstrate how spatial analysis can be used not only to identify geographical hotspots and pandemic dynamics, but also to examine demographic and socioeconomic differences between morbidity and mortality. By linking aggregated data to explanatory factors at an ecological level and using spatial regression analysis, it enables the investigation of disparities between morbidity and mortality even in the absence of individual-level data. Switzerland provides a particularly suitable case study for demonstrating how to harmonise data across different administrative levels, as the epidemics act required municipalities to report influenza cases during the 1918–1920 pandemic. Furthermore, Switzerland remained neutral during World War I and was not affected by war-related excess mortality. Switzerland is also especially relevant for exploring spatial variations at the district level, as political decisions, general healthcare practices, and pandemic mitigation strategies were implemented at the cantonal level or even smaller administrative units ^15^. Moreover, the country’s multilingualism, with German, French, Italian and Romansh regions, creates distinct cultural contexts influenced by neighbouring countries, contributing to regional differences in health status and mortality ^16–18^. These factors make Switzerland an ideal setting both to examine how morbidity and mortality patterns differ and to illustrate our methodological approach.

## 2. The 1918-1920 Pandemic in Switzerland

The 1918-1920 flu pandemic, also known as ‘Spanish flu’, was the most severe influenza pandemic in recorded history. The virus infected approximately 500 million people ^19^, about one-third of the global population at that time. Mortality estimates ranges from in 20-50 million deaths worldwide ^19,20^ . Switzerland was not spared from this pandemic. The first reports of increased influenza cases in Switzerland appeared in May 1918, but it took until the end of June before the country became aware of the pandemic threat. Initial cases were reported among the military, most likely in Basel and western Switzerland and rapidly spread to the civil population ^10,11,21^. Cases were reported sporadically until the end of June, therefore, the end of June to early July is generally considered the actual onset of the pandemic in Switzerland ^11^.

In 1886, Switzerland passed its first epidemic act, which initially covered four diseases - smallpox, cholera, typhoid, and plague - and whose implementation fell under the jurisdiction of the cantons ^22^. A comprehensive revision followed in 1913, making it possible for any infectious disease to be reported ^23^. In the following years, the list of notifiable diseases was gradually expanded. On October 11, 1918, influenza was officially added to this list ^24^. However, the Federal Office of Public Health had already issued a directive on July 6, 1918, requiring weekly reporting of influenza cases ^24^. This recommendation prompted several cantons, including Zurich and Bern, to introduce mandatory reporting as early as July 25, 1918 ^25^ . Other cantons, did not impose a reporting obligation before October 11, 1918 ^24^. The reporting obligation applied to physicians and established the basis for the cantons to report influenza cases to the Federal Office of Public Health on a weekly basis. In some cantons, the weekly figures formed the basis for decisions on measures.

Historical sources show that various non-pharmaceutical interventions were applied during the pandemic. However, there was no nationwide introduction of measures, as the implementation of these measures was the responsibility of the cantons and, at times, even the municipalities ^26^. Previous studies have shown that measures were implemented ^15,27–31^, for example, that summer holidays started earlier and ended later, schools in Valais remained closed from mid-November to early January, while in Bern and Zurich, schools were closed from mid-October to mid-December. Furthermore, public events were banned, theatres were closed, and preaching in churches or visits to hospitals were prohibited.

The effectiveness of these measures was probably decreased by a general strike from November 11-14,1918, right in the peak of the pandemic. Approximately 250,000 workers took part in the strike, opposed by 95,000 soldiers, many of whom were themselves suffering from influenza ^32,33^. As a result, Switzerland experienced a further rise in morbidity and mortality mid-end November 1918.

Switzerland exhibited significant regional differences in geography, population, and economic activity in 1918 ^34^. Switzerland’s cantons (25 cantons in 1918) and its geographical regions (Jura, Swiss Plateau and Alps) are displayed Figure 1.

**Figure 1:**
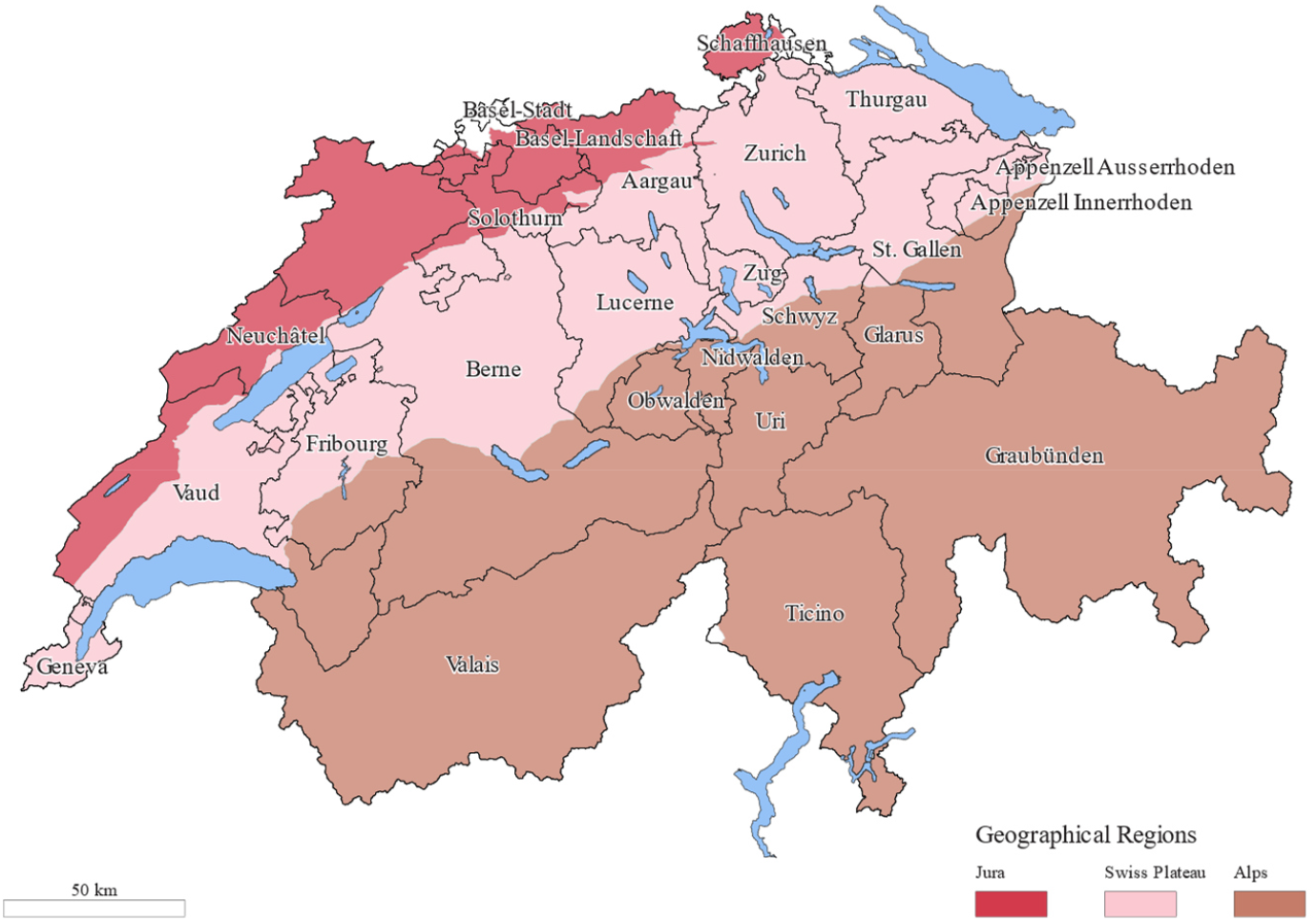
Cantons of Switzerland in 1918 and the geographical regions.

Figure 2 shows the distribution of ecological variables on district level (183 districts in 1918). Urban areas, particularly in the Jura and Swiss Plateau, were densely populated and served as industrial and commercial hubs, with a concentration of multi-family apartment building. Rural areas, especially in the alpine regions, were more sparsely populated, with households being often larger and communities more agrarian in nature. The Alps themselves were characterized by isolated settlements and limited health infrastructure ^29^.

**Figure 2:**
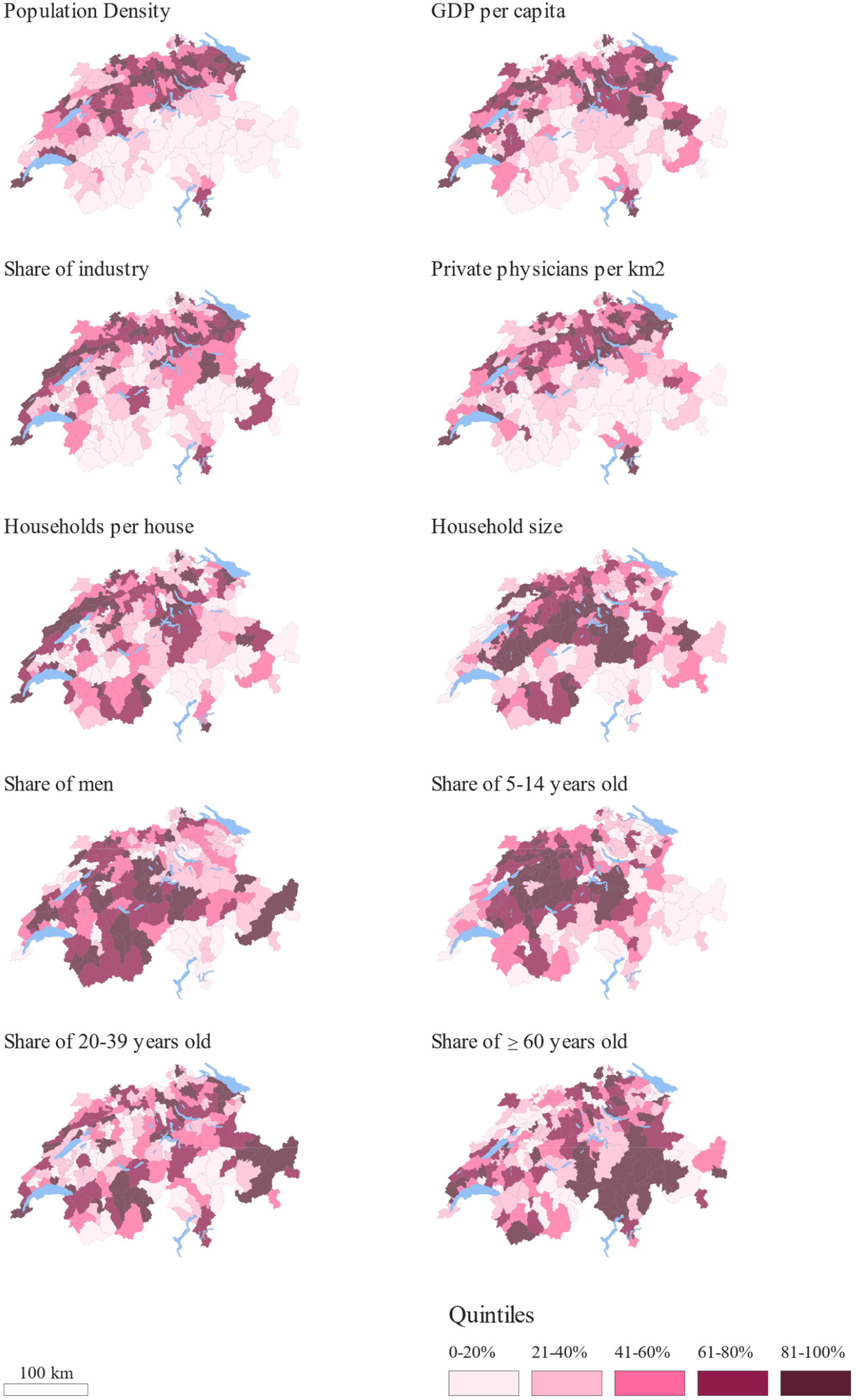
Spatial distribution of ecological variables on district level in 1918.

## 3. Data

### 3.1 Influenza morbidity cases in Switzerland

Since only isolated cases were reported until late June, the onset of the pandemic in Switzerland is generally dated to late June/early July ^11^. We defined three waves: the summer wave from late June to late August 1918 and the second major wave from September 1918 to May 1919. A later third wave, is observed from January to May 1920.

The weekly influenza cases were taken from the Federal Statistical Office’s weekly bulletin on vital statistics (Bulletin des Schweizerischen Gesundheitsamtes), which are freely available online in the Swiss Federal Archives ^35^. The weekly influenza figures were collected from Bulletin 1918 No. 27, covering the week from June 30, 1918 - July 6, 1918 until Bulletin 1919 No. 20, for the week of May 11, 1919 - May 17, 1919, as well as from 1920 Bulletin No. 1, for week of December 28, 1919 - January 3, 1920 to Bulletin No. 18, covering the week of April 25, 1920 - May 1, 1920. In each week, the number of cases were reported for the canton and for the municipalities or at least the district level. Especially during the peak of the pandemic, figures were often only reported at the district level. Therefore, case numbers are analysed only at the district level in this study. In certain weeks, the aggregation of case numbers is inconsistent, with municipal totals not matching district figures and district totals not aligning with cantonal values.

### 3.2 Population at risk

Census data for 1910 and 1920 by district and sex were provided by the Swiss Federal Statistical Office (FSO) ^36,37^. For 1910, additional data were available by 5-year age groups (e.g., 0–4, 5– 9, 10–14, etc.). Annual population estimates by sex and age were linearly interpolated for each district using the 1910 and 1920 census data. Based on this, the age distribution for 1910 was calculated for each district and then applied to estimate the population of each age group for the intervening years.

### 3.3 Ecological determinants

Ecological determinants potentially explaining morbidity and mortality at the district level were collected. Population density was calculated as the total population divided by the district’s surface area. Gross Domestic Product (GDP) per capita was used as defined by Stohr ^34^. Age structure was measured as the share of the population aged 5–14, 20–39, and ≥60 years and the share of men was also calculated for each district. Additional ecological determinants transcribed from the 1920 census ^38^ included the number of private doctors per km^2^, share of industry, mean household size, and households per house. The spatial distributions of the ecological determinates are shown on choropleth maps in Figure 2.

## 4. Harmonizing data on different administrative level

The morbidity data from Switzerland provide an excellent case study to illustrate how morbidity data can be reconstructed and harmonized. The following steps are tailored to the Swiss data but can be applied to other datasets collected across multiple administrative levels. For each week, we checked the internal consistency of case counts across municipalities, districts, and cantons, from the lowest to the highest administrative level. Reported case counts were harmonized using a stepwise adjustment procedure, as shown in Figure 3. This procedure ensures that municipality totals never fall below district totals, district totals never fall below canton totals, and any excess at the cantonal level is fairly reallocated while preserving the minimum reported values. Furthermore, this method ensures that the maximum total number of cases is preserved, as all administrative levels are included and any underreporting at a given level is compensated through the harmonization process.

**Figure 3:**
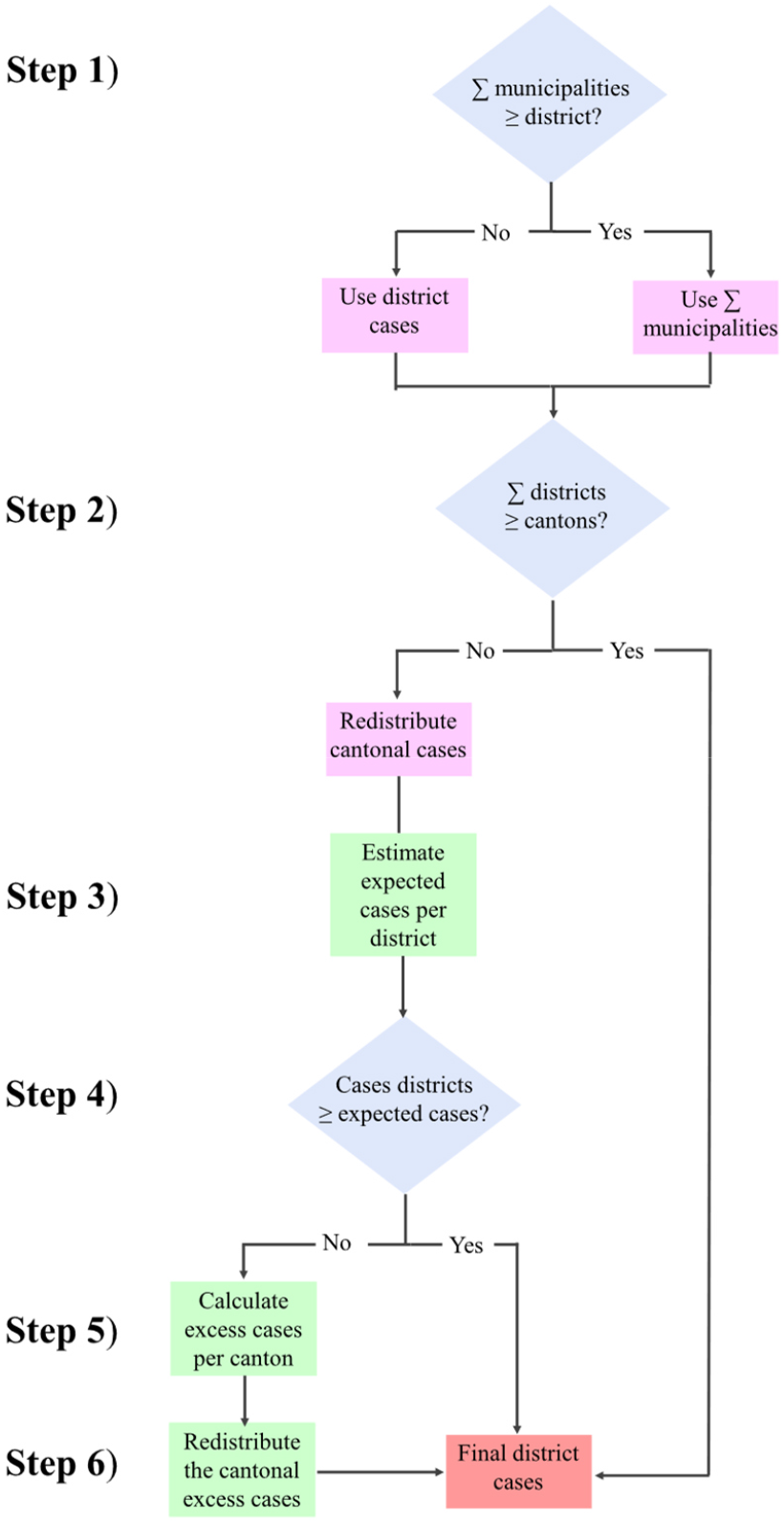
Flowchart of the stepwise adjustment procedure to estimate the district cases

The following section describes the steps used to compile case numbers at the district level. **Step 1) Compare municipality vs. district cases:** The reported number of cases *C*_*d,w*_ for each district *d* and in each week *w* is compared with the sum of the cases of the municipalities *C*_*m,w*_ in each district *d* and in each week *w*. If the sum of the cases in the municipalities is larger than the numbers of cases in each district then the sum of the cases of the municipalities is taken as number of cases for the district 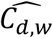, otherwise the cases in each districts. This procedure ensures consistency across municipality and district level.

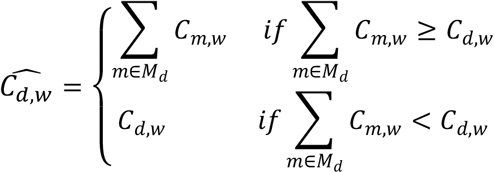

**Step 2) Compare district vs. cantonal cases:** If the sum of the cases in each district for each canton 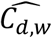 is larger or equal than the reported cantonal cases *C*_*c,w*_ the adjusted 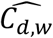 is taken as final number of cases in the districts 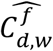, otherwise the final 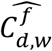 is estimated further in Step 3). This procedure ensures consistency across district and cantonal level.

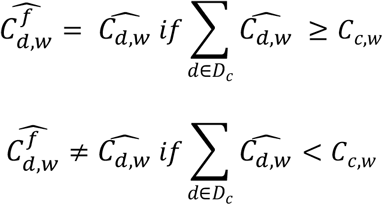

**Step 3) Estimate expected cases per district**: The expected cases for each district *E*_*d,w*_ given the cases in each canton *C*_*c,w*_ and the total population in each district *pop*_*d,w*_ were estimated by

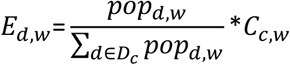

**Step 4) Compare adjusted cases vs expected cases per district:** The adjusted cases for each district and week 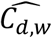 were compared with the expected number of cases in each district and week *E*_*d,w*_. If 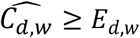 then 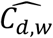 was the final number of cases for each district 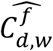. For all districts for which 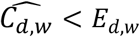, the final 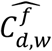 is estimated further in Step 5).

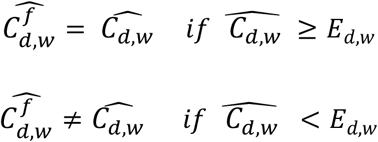

**Step 5) Calculate excess cases per canton:** For each canton and in each week where 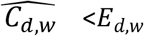 the excess cases *R*_*c,w*_ are calculated

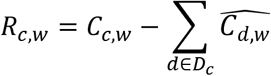

**Step 6) Redistribute the cantonal excess cases**: For all districts for which 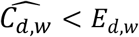 the excess cases *R*_*c,w*_ was redistributed across districts in proportion to their population shares and summed up to 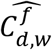. This ensures that no district was assigned fewer cases than its original report.

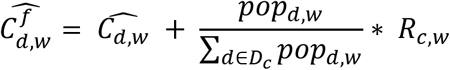

### 4.1 Quality check

District data was missing in 13%, and in 12% the district case numbers are adjusted upward due to higher canton-level figures. The canton of Fribourg is particularly affected, the very first weeks of the pandemic 1918 and some weeks in 1920. Otherwise, missing or underestimated district values were randomly distributed across weeks and districts. For redistributing excess canton cases, the total population of each district was used. A sensitivity analysis was also performed using the population aged 20–39, based on the assumption that this age group was primarily affected by the pandemic, with districts having a higher share of 20–39-year-olds receiving greater weight. As the sensitivity analysis yielded very similar results (not shown), the total population was retained for the estimation.

A total of 892,865 cases were reported on a population just over 4 million people. Some researcher suggested an incidence of approximately 2 million cases in Switzerland (corresponding to about 50–60% of the population)^10,39^. However, the course of the morbidity data for whole Switzerland confirms the documented pattern of 1918-1920 mortality studies in Switzerland with of three waves (Figure 4) ^10,28,40,41^: a first mild wave in the summer of 1918 (total number of cases: 101,406), a severe wave in the fall of 1918 (total number of cases 649,662), where the renewed rise in cases after the national strike is clearly visible, and a real mild wave at the beginning of 1919. In addition, there was a later, third wave in the first quarter of 1920 (total number of cases: 141,797).

**Figure 4:**
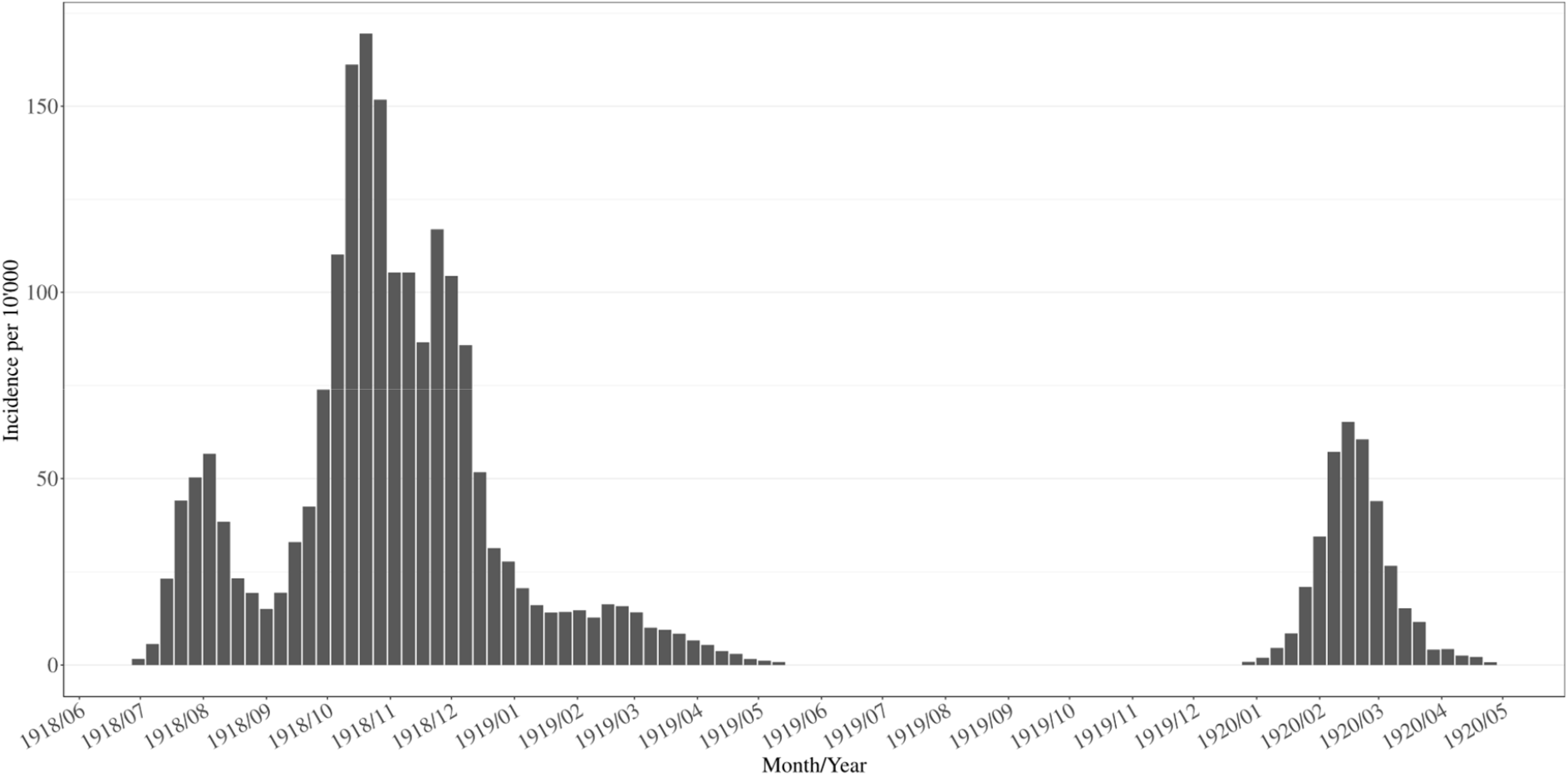
Weekly incidence for whole Switzerland.

When comparing monthly incidence rate with the monthly excess mortality in Switzerland published in Staub et al. ^40^ (Figure 5), the trends appear very similar. Only in September is the excess mortality lower and in November notable higher, which may be due to underreporting, but could also reflect deaths among individuals who contracted the disease in October, thereby illustrating the time lag between disease onset and mortality.

**Figure 5:**
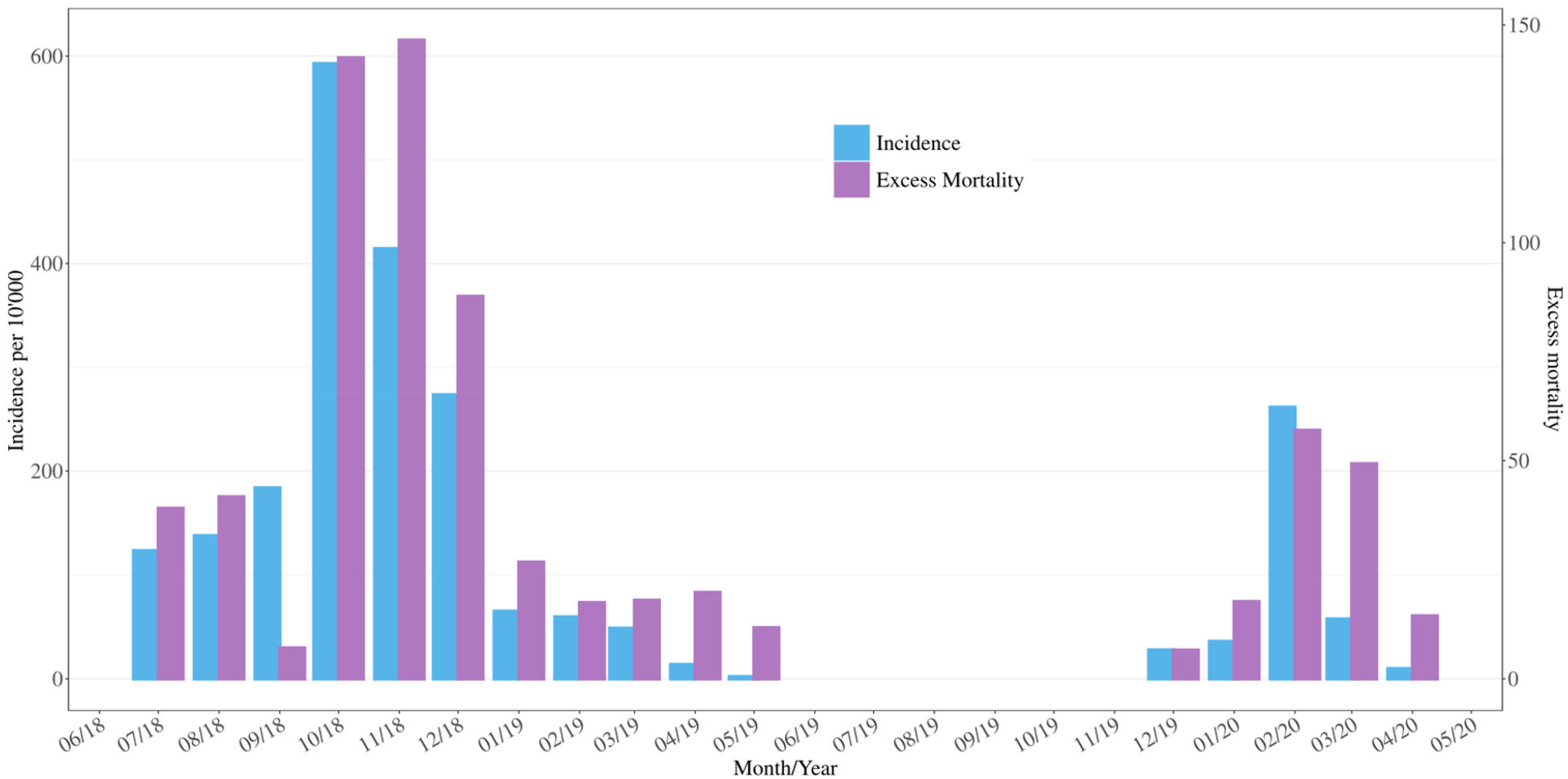
Comparison of monthly incidence and excess mortality in Switzerland. Excess mortality values are from Staub et al. ^40^

Even though case numbers are probably underestimated, the majority of cantons display temporal patterns comparable to those observed in mortality data allowing for relative spatial comparisons. In Figure 6, the course is displayed for each canton. Most of the cantons show the expected pattern. However, Appenzell Innerrhoden (AI) recorded no cases of influenza between August 1918 and May 1919. which is very unlikely, as Appenzell Innerrhoden also experienced excess mortality in 1918 ^5^ . However, this does not affect the overall analysis, as it was the smallest canton in 1918, with only 14,500 residents and just 351 deaths that year. Elsewhere, the development of cases in Fribourg (FR), Glarus (GL), and Valais (VS) appears somewhat irregular, which could be related to delayed reporting in the second strong wave. However, since the later analysis focuses on waves rather than specific weeks, this does not pose an issue. Moreover, the overall pattern - a mild first wave, a severe second wave, a very mild 1919 wave, and an additional wave in 1920 - also aligns with the course observed in mortality studies in Switzerland.

**Figure 6:**
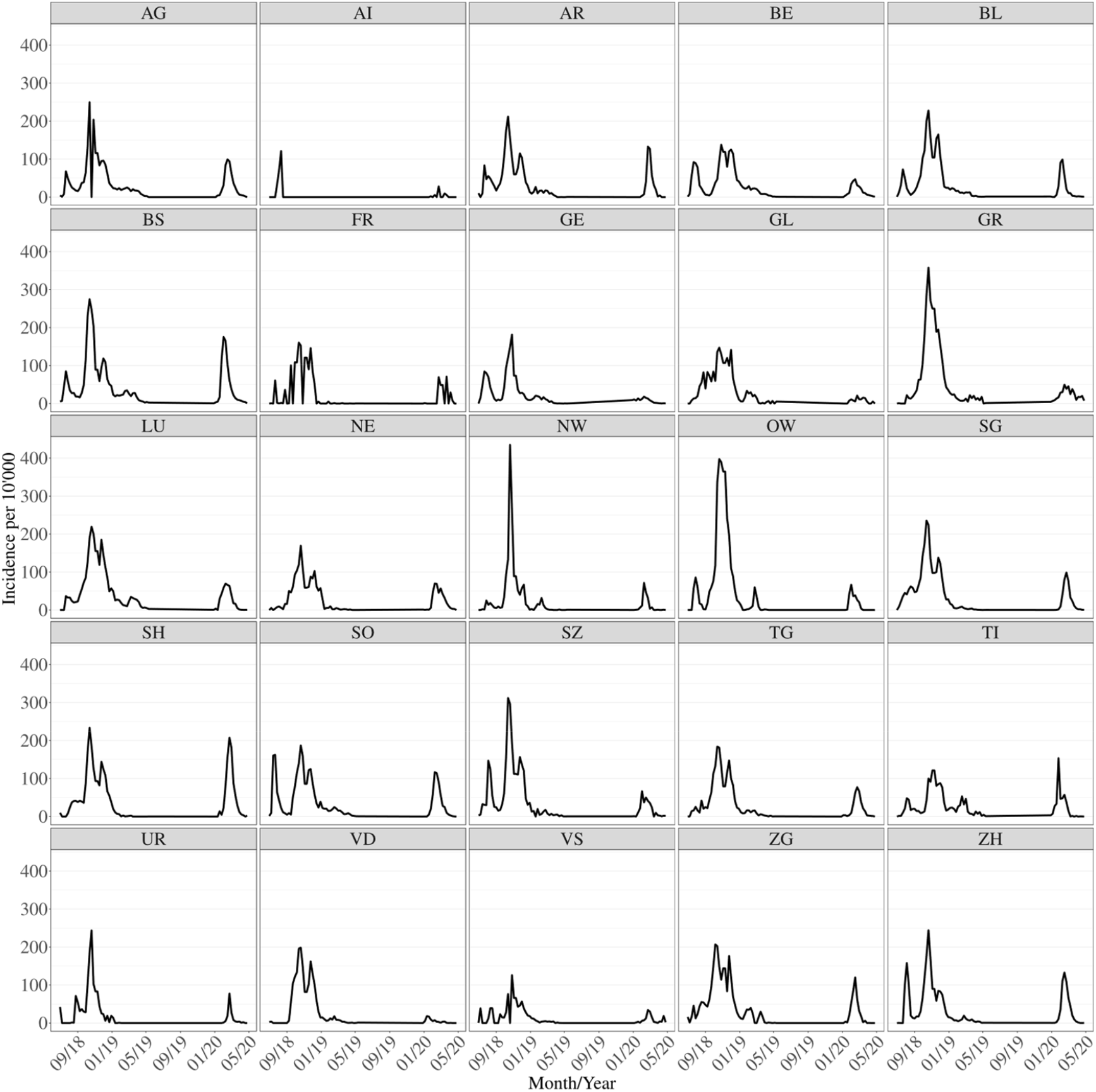
Weekly incidence for each canton.

Examining the differences between incidence and excess mortality by district for 1918 (Figure 7) shows that, in most districts, incidence and excess mortality align closely. Notable discrepancies are observed in Valais, where the incidence rate is considerably lower than excess mortality (see also the excess mortality map, Supplementary Figure S2), which is likely attributable to the limited availability of physicians and reflect an underreporting of cases here. Conversely, high morbidity does not necessarily translate into high mortality.

**Figure 7:**
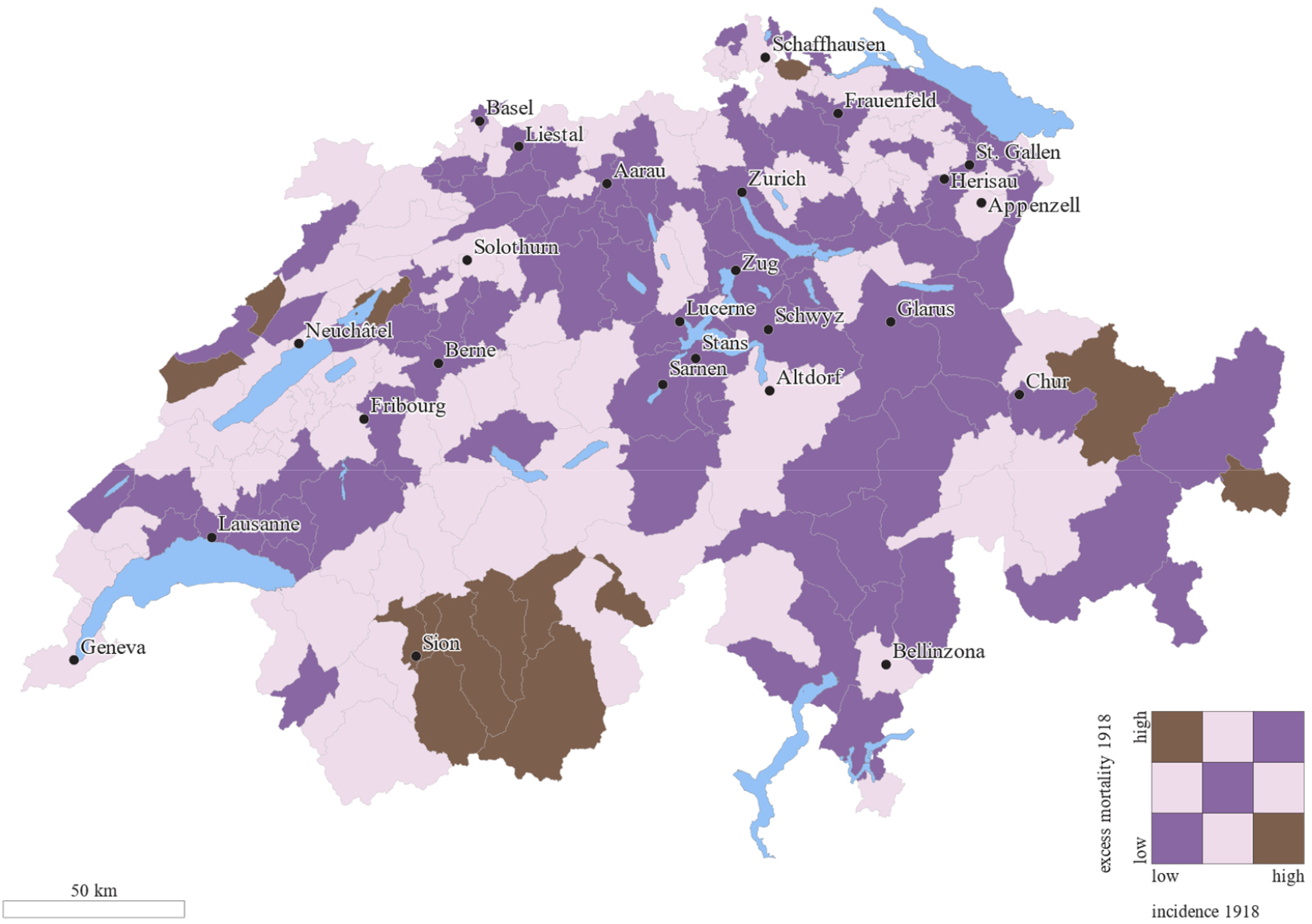
Areas shown in brown represent weak correspondence between incidence and excess mortality, whereas dark purple indicates a strong correspondence. The relationship may be positive in either direction, reflecting both low–low and high–high combinations of incidence and excess mortality.

Based on the presented data explorations, we conclude that the quality of the morbidity data is sufficient to make a meaningful contribution to understanding the 1918–1920 influenza pandemic in Switzerland after the implementation of a stepwise adjustment procedure to reconcile inconsistencies in reported case numbers across different administrative levels. In the next section we will use these harmonized morbidity data to study differences in determinants of morbidity and mortality by using spatial analysis methods.

## 5. Using harmonized morbidity data in a spatial context – the Swiss case study

### 5.1 Statistical methods

The incidence rates per district were calculated per 10,000 inhabitants. The monthly population per district was estimated by linear interpolation between the annual population figures. Monthly incidence rates for each of the 183 districts is displayed in choropleth maps using quartiles. Getis-Ord Gi* statistic ^42^ is applied to identify clusters of districts with higher or lower incidence rates. The Getis-Ord Gi* statistic is expressed as a z-value: values greater than 2 indicate significantly higher incidence rates, while values less than -2 indicate significantly lower rates compared to Switzerland overall. A nearest-neighbour approach was used, with five neighbours providing the best model fit. The geometries of the districts that formed the basis of the spatial analysis were obtained from the Swiss Federal Statistical Office ^43^. Additionally, datasets from the Federal Office of Topography (swisstopo) were used to depict Switzerland’s geographical regions and topographic relief ^44,45^.

The spatial diffusion of each of the three waves was estimated using the maximum incidence rate in each district in each of the three waves. Especially at the beginning of the pandemic, the first cases were not reported, making it difficult to trace the spread based on initial notifications. Assuming a relatively constant number of physicians diligently reporting each week, and a comparable baseline reproduction number across Switzerland, we consider the peak of each wave in every district to be a reliable measure of the diffusion process.

An exploratory analysis was conducted to examine the relationship between the described ecological variables and the incidence rate for each wave. All explanatory variables were z-transformed for comparability. A Geographically Weighted Regression (GWR) was used ^46^, which extends ordinary least squares (OLS) by allowing relationships between independent and dependent variables to vary geographically, generating localized models to assess spatial variation in coefficients. To overcome the issue of outliers and extreme values a robust GWR is applied. Because the ecological variables were highly intercorrelated (see the correlation matrix in Supplementary Figure S4), each was analysed in a separate univariable regression to estimate its unadjusted association with the outcome. The model can be specified as:

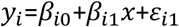

Where *y*_*i*_ denotes is the dependent variable, namely incidence rate, at location *i*, and *β*_*i0*_ the intercept, *β*_*i*1_*x* the coefficient representing the change in in *y*_*i*_ due to a one-unit increase in *x*, and ε_*i*1_ the error term at location *i*. The parameters in GWR are estimated using weighted least squares, with a diagonal weighting matrix *w*_*ij*_. A gaussian kernel density weighted function was used to determine the weights:

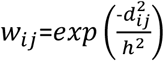

where *d*_*ij*_ is the distance between location *i* and *j. h* is the bandwidth. The optimal bandwidth (distance of neighbours used for each local regression) was found via an adjusted Akaike Information Criterion (AICc) and an adaptive bandwidth with a fixed number of neighbours was used ^47^. Besides the local-specific coefficients for each independent variable, which are shown on choropleth maps, the median coefficients across all 183 districts were calculated and 95% bias-corrected and accelerated (BCa) bootstrap confidence intervals (CIs) ^48^ were estimated by using 2,000 samples.

To compare incidence and mortality, we calculated excess mortality using all-cause deaths, as district-level data on influenza-specific deaths are unavailable. Excess mortality indicates how many more individuals than expected died in each district during the pandemic. We estimated excess mortality for 1918 by using a Bayesian spatial model for disease mapping, following the approach used in a previous paper on excess mortality ^5^. Unlike that study, however, we included all 183 districts rather than harmonizing the data to 130 districts. Since overall mortality data were not available at weekly or monthly level, excess mortality could only be calculated at annual level. Since excess mortality in Switzerland was low in 1919 and 1920, we restrict the comparison to 1918, the most severe pandemic year with markedly elevated excess mortality ^5^. To investigate how ecological determinants relate to the spatial distribution of incidence compared to the excess mortality, we used the z-values from the G* analysis of incidence rates as the outcome variable. The excess mortality was also transformed into z-scores. A robust univariate linear regression model was employed.

All statistical analyses were performed using R Version 4.5.0 ^49^. The R package GWmodel for the GWR regression^50^. Getis-Ord Gi* statistic and map creation were carried out using ArcGIS Pro, version 3.5.2^51^. The data is publicly available via zenodo https://doi.org/10.5281/zenodo.17508352 and the code via GitHub https://github.com/KaMatthes/Spatial_temporal_morbidity_1918

### 5.2 Results of the Swiss case study

Figure 8 shows the incidence rate per month in each district and Figure 9 the results of the Getis-Ord Gi* analysis. A z-score below 0 means that the incidence in these regions was lower than the Swiss average, which was in most cases still elevated. In July, a pandemic hotspot emerged in North-western Switzerland (Jura). By August, the epidemic focus had shifted further eastward, although parts of the northern region, particularly the Jura, remained affected. The entire canton of Vaud reported no cases in August. In September, the pandemic advanced further east into Grisons (Graubünden), where distinct hotspots became apparent, while incidence in the rest of Switzerland began to decline. During October and November, the incidence increased substantially, with hotspots relocating toward central Switzerland.

**Figure 8:**
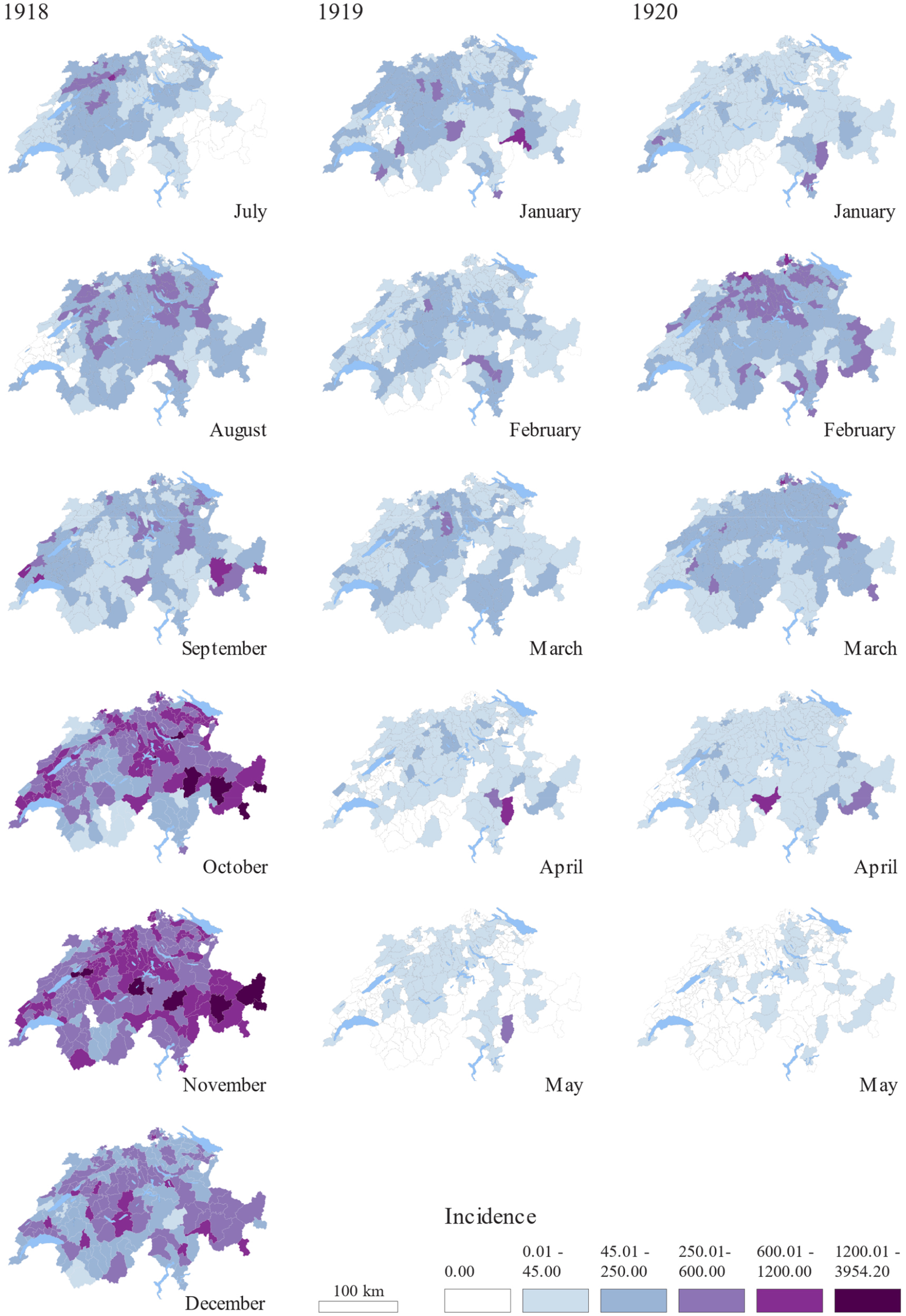
Monthly incidence rates in each district

**Figure 9:**
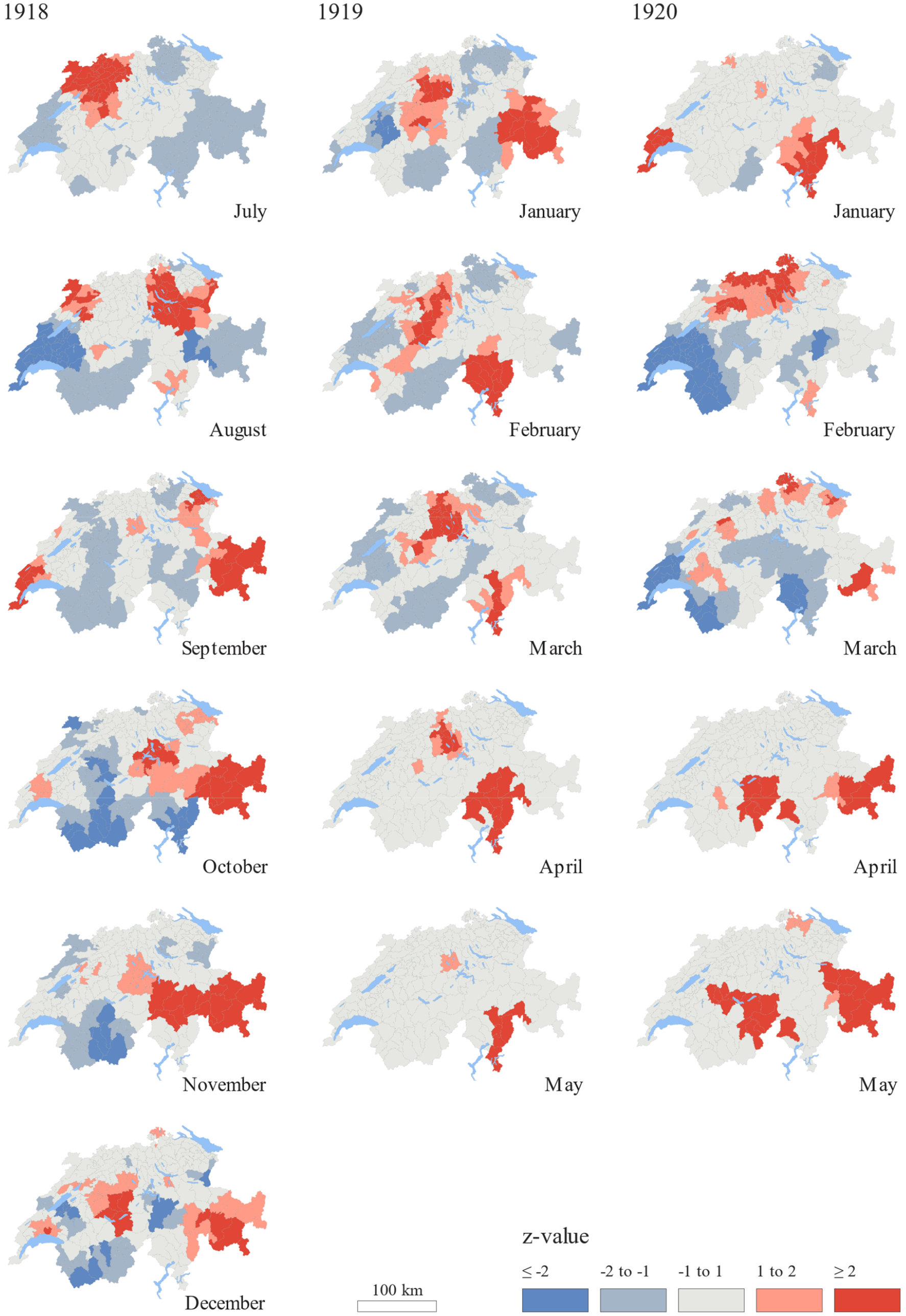
Monthly results of the Getis-Ord Gi* analysis Higher z-values indicate greater intensity of clustering and the direction (positive (red color) or negative (blue color)) indicates a cluster of high or low incidence rate.

From December 1918 to May 1919, infection rates gradually declined throughout Switzerland. However, southern Switzerland (the canton of Ticino) continued to record elevated infection rates in 1919. While the pandemic had largely subsided in most parts of the country, several districts in Ticino continued to report significant numbers of cases. In January 1920, the first hotspot seems to reappear in Geneva in the west, and another one in Ticino in the south of Switzerland. In February, the northern part of the country is particularly affected. From March onward, case numbers gradually decline across all cantons. In April and May, some hotspots remain in the east of Switzerland, in Grisons, and in the south, although case numbers decreased here as well and are low in May.

Figure 10 displays the spatial diffusion for each wave by showing when each district hit its maximum incidence rate during the three observation periods. In the first wave of 1918, from July to August, it becomes very clear how the first peak was reached in Geneva in western Switzerland, then spread from west to northeast (Jura and Swiss Plateau), with the southern part (Southern Alps) of the country being affected last. In the second wave, there is no obvious spatial diffusion; the peaks in the districts appear rather randomly. What does become apparent, however, is that Ticino in southern Switzerland was affected later, some districts even in 1919. In 1920, the spread began again in western Switzerland, but at the same time, peak values were already observed in Ticino and Grisons (in the east). Very roughly, the progression can be seen from the west toward central Switzerland and the northeast, and later to Valais in the south.

**Figure 10:**
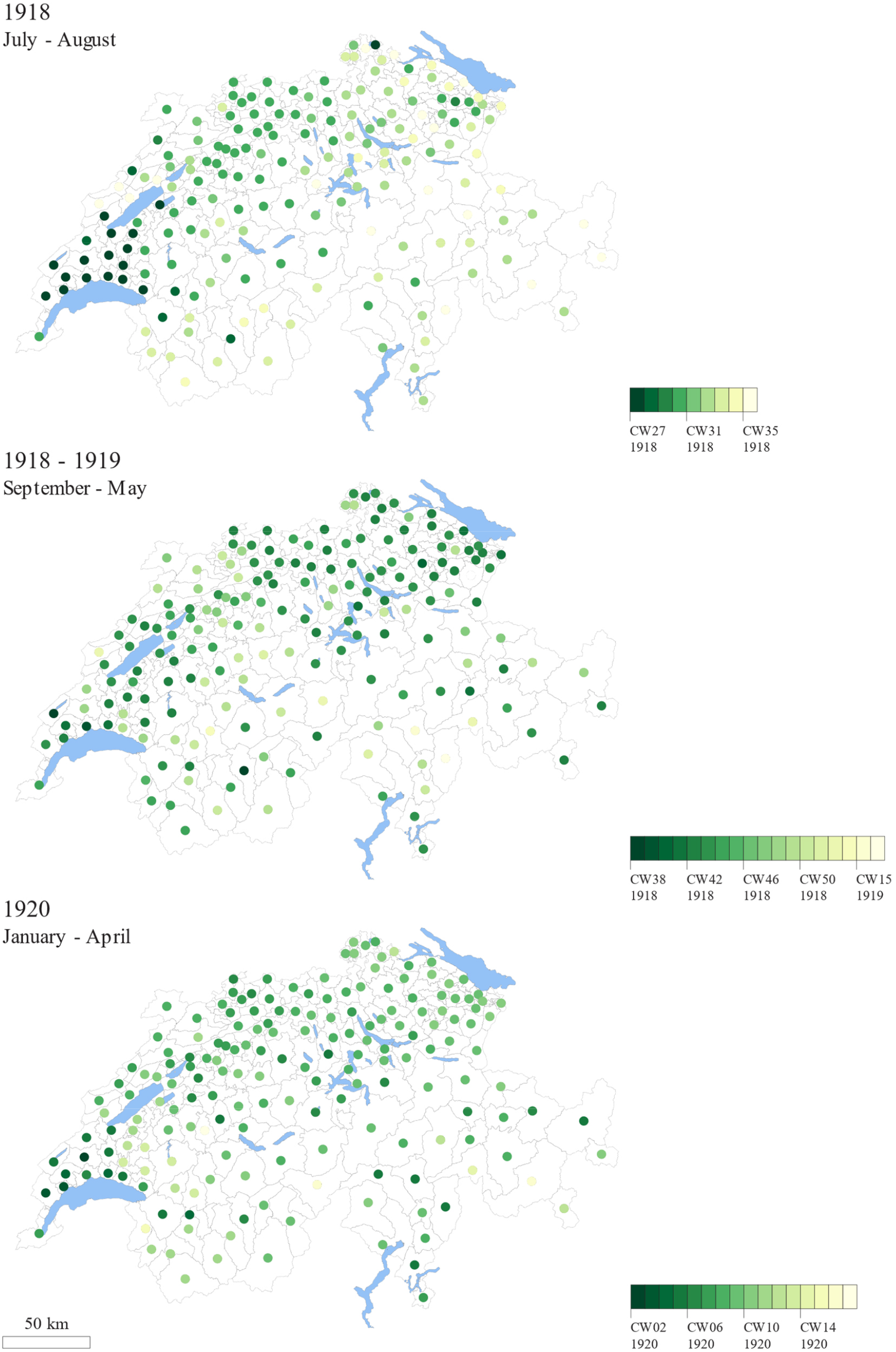
Spatial diffusion using the maximum incidence rate in each district in each of the three waves

Figure 11 presents the association between the median of ecological factors and morbidity of the 183 districts for each wave and explanatory variable (detailed results in Supplementary Table S1).

**Figure 11:**
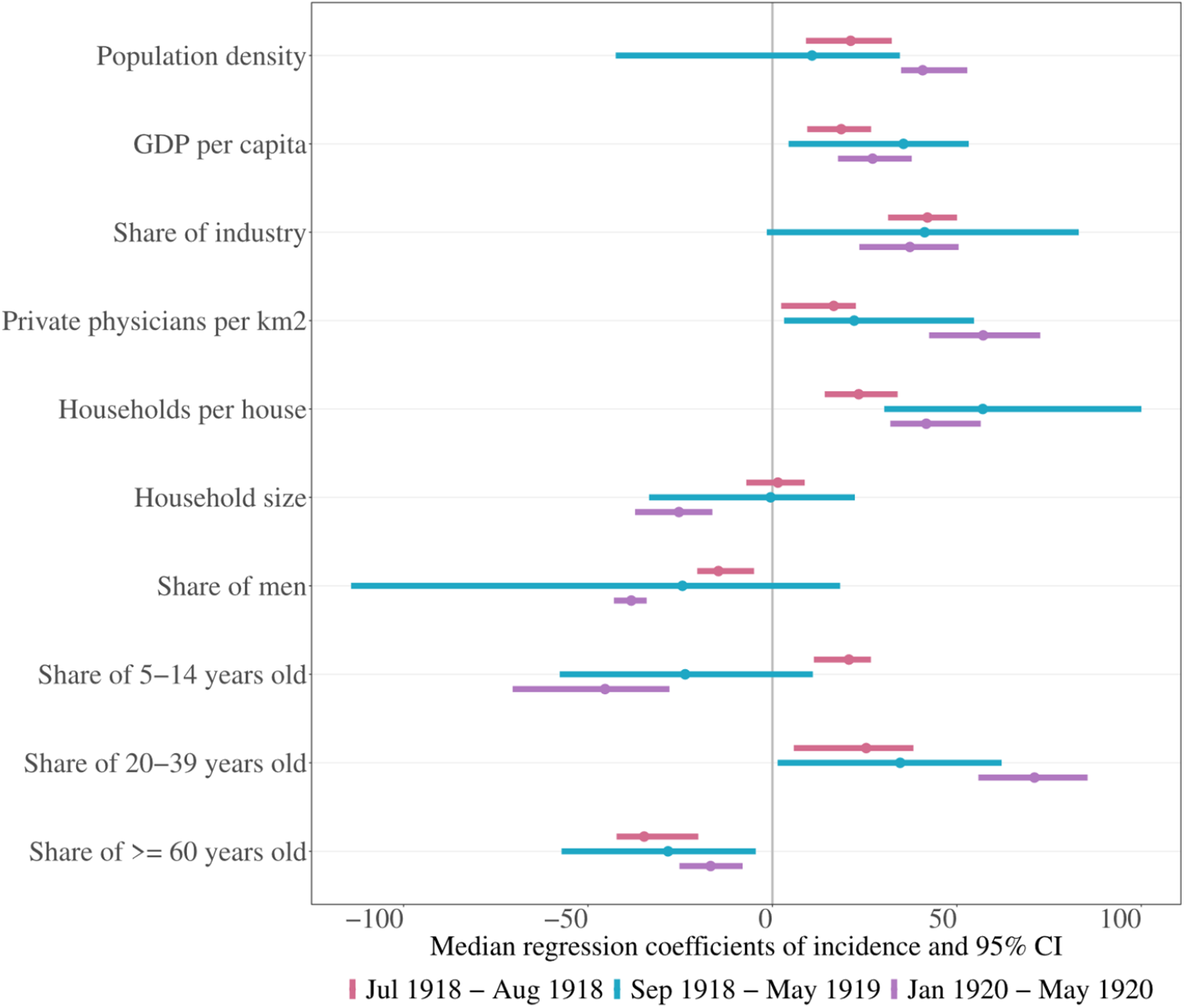
Median regression coefficient of incidence and 95% confidence intervals (CI) from the robust GWR. Each determinate is showing one model (univariable)

Population density, GDP per capita, the share of industry, the number of private physicians per km^2^, and the number of households per house are all associated with higher incidence rates across all waves. The share of men is associated with lower incidence rates in all waves, although in the second wave (September 1918 – May 1919) this relationship appears only as a trend. The proportion of children aged 5–14 years is associated with higher incidence rate during the first wave (July 1918 – August 1918), but not in the second and third waves, where incidence is lower, though only a trend is visible in the second wave. A higher share of individuals aged 20–39 years is associated with a higher incidence rate, while a higher share of those aged 60 years and older is associated with a lower incidence rate. However, in the second wave the confidence intervals are much wider than in the other waves, indicating that the influenza had spread throughout the entire country and that many districts experienced very high incidence rates.

The corresponding choropleth maps of the regression coefficients are shown in Figure 12-14. When examining the regression coefficients of the individual districts, local differences become apparent. The share of men and the share of individuals aged 20–39 play an important role during the first wave (Figure 12), particularly in the northwest, where the highest incidence was also observed. The share of people aged ≥60 years is associated with lower incidence rates across most of Switzerland; however, there are districts in Valais where a higher share of this age group corresponds to higher incidence. Population density, GDP per capita, the share of industry, and the number of households per house appear to be less strongly associated with high incidence in western Switzerland than in the rest of the country. In contrast, factors such as household size and the share of children aged 4–15 years seem to be more influential there.

**Figure 12:**
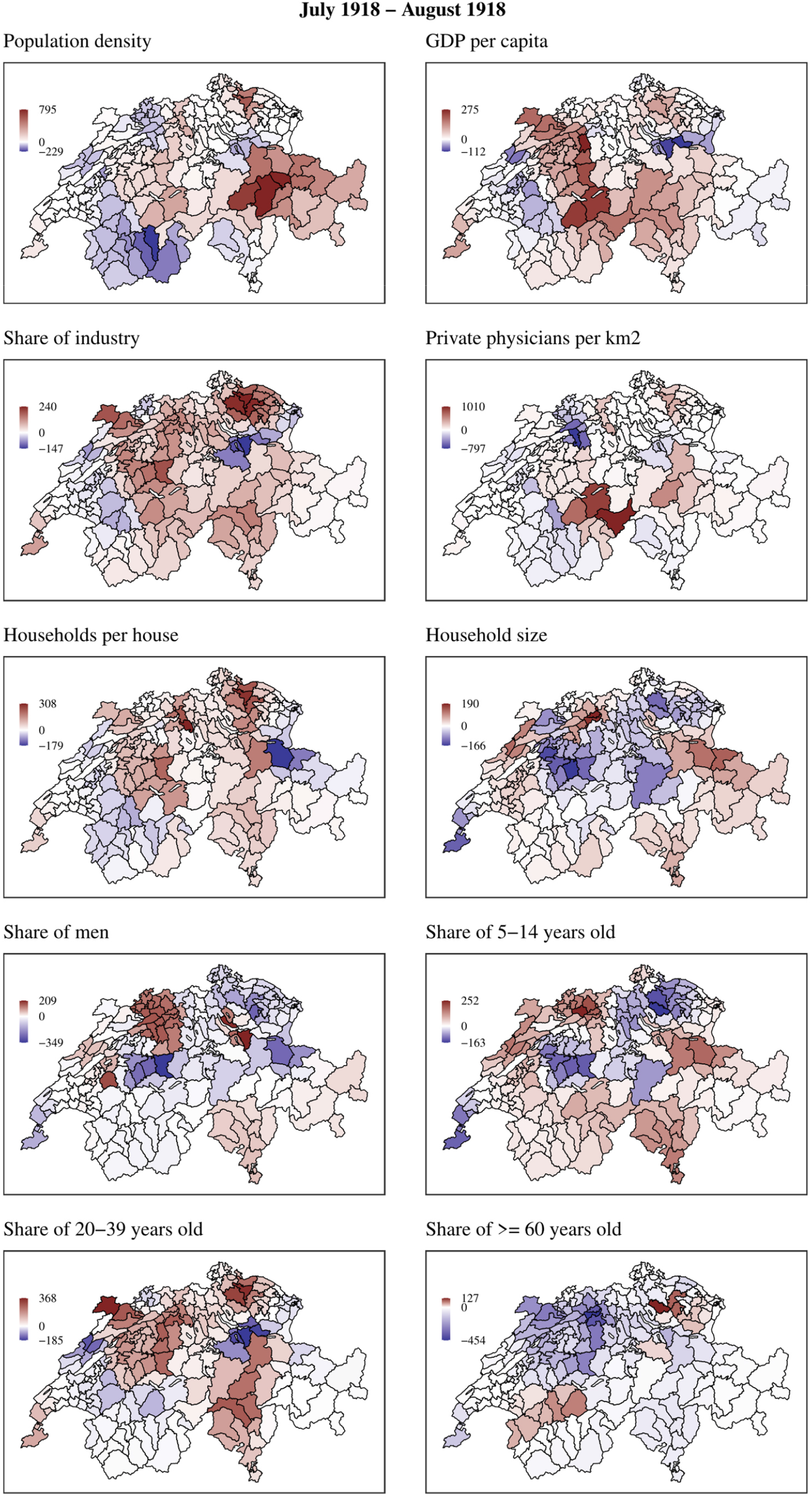
Regression coefficients of the GWR model for the first wave (July–August 1918). Red indicates a positive association between higher incidence rates and higher values of the determinant, while blue indicates a negative association.

In the second wave (Figure 13), the pattern looks different. For example, when examining the age groups and their influence on incidence, a higher share of children aged 4–15 years is still associated with higher incidence rates in eastern Switzerland (Grisons) and in Ticino. In contrast, the 20–39 age group is particularly associated with higher incidence on the northern side of the Alps, in the Swiss Plateau, and in the northeast. The ≥60 age group is associated with higher incidence in various parts of Switzerland. The share of men also appears to be linked to higher incidence in the Jura and Swiss plateau and in Ticino.

**Figure 13:**
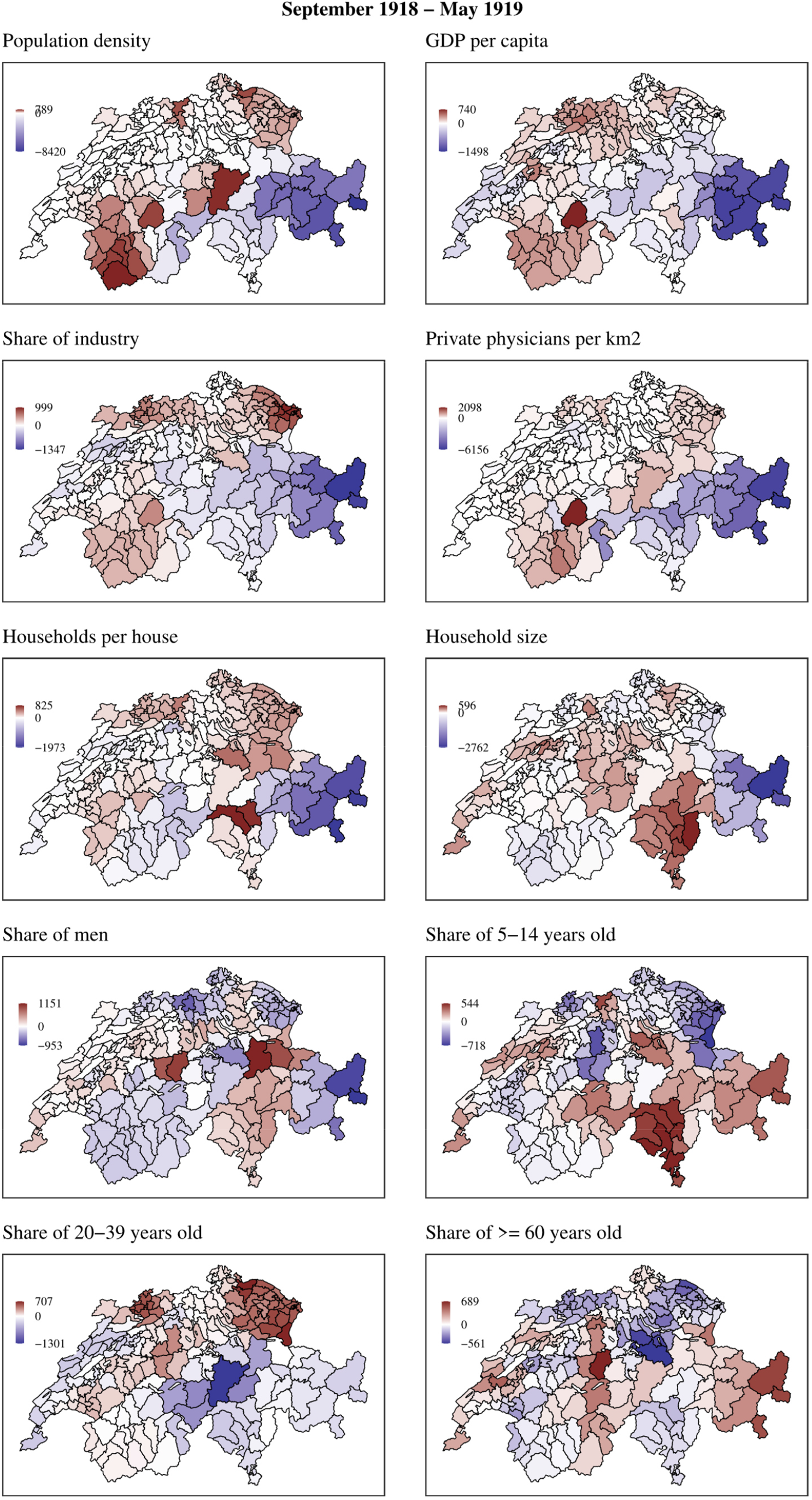
Regression coefficients of the GWR model for the second wave (September 1918 – May 1919). Red indicates a positive association between higher incidence rates and higher values of the determinant, while blue indicates a negative association.

From January to May 1920 (Figure 14), the share of individuals aged 20–39 is particularly associated with higher incidence in the German-speaking part of Switzerland, whereas the share of those aged ≥60 years is more strongly associated with higher incidence in the French-speaking part.

**Figure 14:**
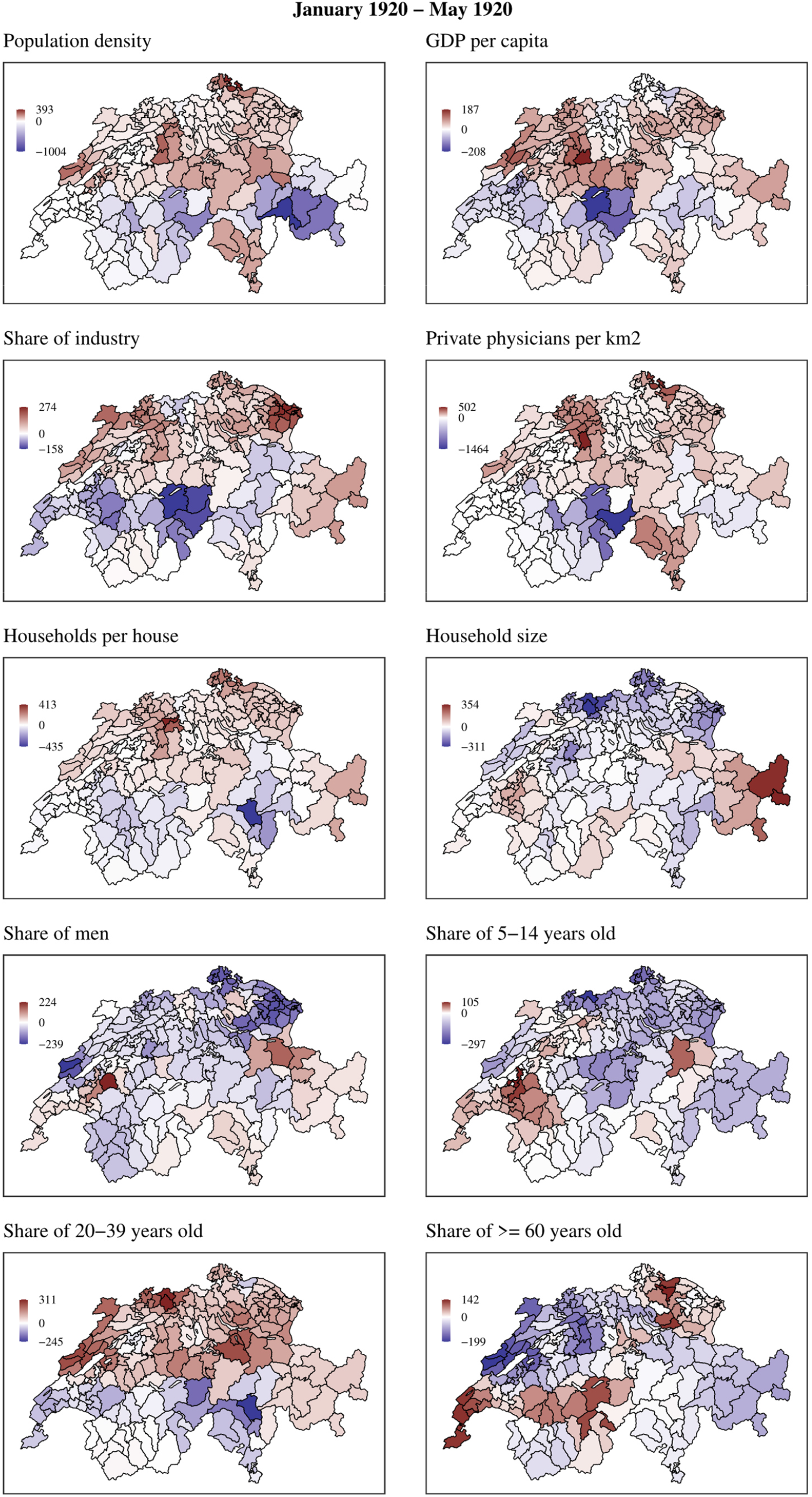
Regression coefficients of the GWR model for the second wave (January 1920 – May 1920). Red indicates a positive association between higher incidence rates and higher values of the determinant, while blue indicates a negative association.

Figure 15 shows results of the robust linear regression for the annual incidence rate and excess mortality for 1918 (detailed results in Supplementary Table S2). Incidence rates were higher in districts with a higher GDP per capita and a larger industrial sector, but these factors were not associated with a higher mortality. Interestingly, the proportion of men showed a negative trend with incidence but a positive trend with excess mortality. All other explanatory factors show similar pattern between morbidity and mortality.

**Figure 15:**
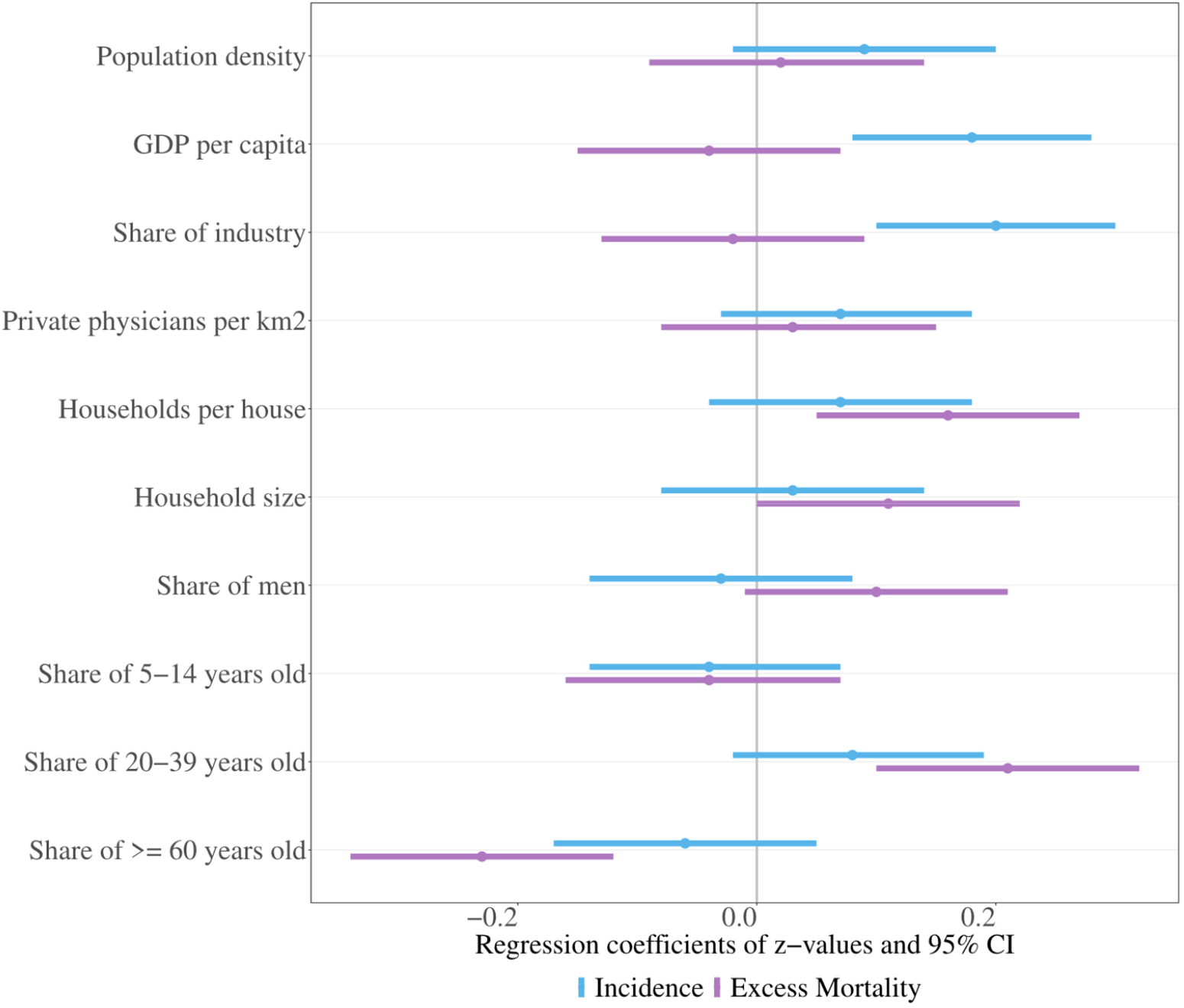
Regression coefficient and 95% confidence interval (CI) of the robust linear regression models. Each determinate is showing one model (univariable).

### 5.3 Discussion of the Swiss case study

In the Swiss case study, we examined spatial patterns and differences in explanatory factors of morbidity data and demonstrated that, despite the high likelihood of underreporting, incidence data provide valuable insights into the dynamics of pandemics. This is particularly relevant as certain determinants of incidence differ from those associated with mortality. Our findings indicate that the quality of the morbidity data is sufficient to allow for meaningful analyses of spatiotemporal dynamics and determinants. Nevertheless, we caution that absolute case numbers should not be interpreted in isolation. Instead, the value of these data lies in their relative interpretation, revealing temporal dynamics and differences across districts. In this regard, incidence patterns can still be considered robust and epidemiologically informative, even if exact case counts remain uncertain, as is also the case in more recent pandemics.

In the first wave, the influenza dynamic is clearly visible, beginning in the west and progressing eastward and then southward. This pattern reflects the fact that troops arriving from France in the west were affected first. During the troop mobilization in western Switzerland at the beginning of July, 13 recruit schools were severely impacted by the flu, with the northern Jura region and Neuchâtel being the hardest hit ^21^. The monthly morbidity data also mirror the monthly spatial mortality data reported by Sonderegger ^11^. The GWR of the first wave confirms a higher incidence rate among men aged 20–39 in western Switzerland, likely reflecting the young male military population. In the second wave, Switzerland was overall affected, but especially high in the cantons of Nidwalden (NW), Obwalden (OW) and Grisons (GR) in central and east Switzerland, which is in line with the spatial mortality results of Sonderegger ^10,11^, showing a high influenza mortality in the second wave for these cantons. 1920 was not considered by Sonderegger and cannot be compared.

Across all waves, higher incidence rates were positively associated with population density, GDP per capita, the share of industry, and the number of private physicians per km^2^. Examining the distribution of these variables in Switzerland (Figure 2) reveals a similar pattern across the country, with the highest concentrations observed in the Jura and the Swiss Plateau The correlation matrix (Supplementary Figure S1) further shows that these variables are highly correlated, suggesting that they likely capture the differences between rural and urban districts. Regions with a higher density of multi-family apartments were associated with higher incidence rates, whereas the size of households played a lesser role. However, the number of multi-family apartments also reflects regions with a higher proportion of industry, except in Valais where the proportion of industry is low, yet the density of multi-family apartments is relatively high. Household size tends to be larger in rural areas of Switzerland, which are characterised by lower levels of industrialisation. We only have information on household size, but not on the average living space per household in each district. Mamelund showed that smaller living space (used as a proxy for lower socioeconomic status) was associated with higher incidence, particularly during the first summer wave ^52,53^.

During the summer wave, districts with a higher proportion of children of school age (5–14 years) were particularly affected, confirming findings from survey studies conducted in the United States ^4^. The proportion of individuals aged 20–39 years was associated with higher incidence rates across all waves; in the first wave, this was likely related to men serving in the military. The proportion of individuals aged ≥60 years was associated with lower incidence in all waves, reflecting the lower mortality observed in this age group.

During the second wave, the confidence intervals were substantially wider than in the other waves, indicating that influenza had spread extensively across the entire country and that many districts experienced very high incidence rates. This highlights the widespread nature of the outbreak during this period in Switzerland, especially in November 1918, compared with the more localized patterns observed in other waves.

The present study also reflects the paradox that, while more women were reported to be infected, more men died ^1,54^. Since most studies are based on mortality data, a higher risk of disease from the 1918-1920 influenza is often anticipated for men. Yet, women appear to have been more affected by the pandemic, but not by the mortality. This finding is probably even stronger than reported, since the analysis relies on cases reported by physicians, which in the first wave had a very high coverage for the troops.

GDP per capita and the share of industry also show opposite patterns for morbidity and mortality. This may indicate that districts with individuals with higher SES were more likely to visit a physician (as only physicians reported cases, leading probably to a selection bias toward higher SES), but had a lower risk of dying from influenza. Socioeconomic differences in mortality have already been demonstrated in several studies on the 1918 pandemic ^55–61^.

The district-level ecological variables provide reasonable approximations and offer a valuable overview of spatial morbidity patterns. However, many of the ecological variables are correlated with each other, reflecting overlapping dimensions of the same underlying conditions (e.g., the strong influence of population size, such as rural versus urban contexts). The results should therefore be interpreted with caution and regarded as exploratory. Despite some resistance from the cantons to reporting influenza, and the fact that several cantons refrained from mandatory reporting during the first wave ^62^, undoubtedly leading to an underreporting of cases, particularly in Valais which is likely attributable to the limited availability of physicians ^63^. Nevertheless, our analysis shows that the wave-like pattern of the pandemic was evident across all cantons. Moreover, we are unable to account for specific public health measures, as district-level data are not available. Although there were no nationwide regulations in Switzerland, evidence from other studies conducted in various Swiss regions indicates that such measures were implemented in 1918 and 1920 ^15,27–31^.

## 6. Conclusion

We demonstrate that morbidity data can make a significant contribution to understanding the 1918–1920 influenza pandemic in Switzerland by complementing existing mortality-focused research with a morbidity perspective. Our results show that the factors shaping morbidity patterns may differ from those influencing mortality, underscoring the importance of examining both dimensions to achieve a more comprehensive understanding of pandemic dynamics. Although morbidity data are often considered less reliable, they can still provide valuable insights into the course and impact of a pandemic. While caution is necessary when interpreting absolute case numbers, we argue that relative analyses, particularly when incorporating data from different geographic and administrative levels, remain highly informative. We showed that morbidity data can be successfully harmonized through a stepwise procedure that preserves the highest plausible case counts. Moreover, the use of spatial data allows not only the identification of pandemic dynamics and hotspots, but also the exploration of potential determinants, even in the absence of individual-level or individual-level data without detailed demographic and socioeconomic information. Hence, this paper shows that historical demographers and historical epidemiologists can and should utilize morbidity data to study pandemics, rather than dismissing it outright due to perceived data quality problems.

## Data Availability

All data produced are available online at zenodo and GitHub

https://github.com/KaMatthes/Spatial_temporal_morbidity_1918

https://doi.org/10.5281/zenodo.17508352

## Author statements

### Ethical statement

Ethics approval was not required for the reuse of these aggregated and publicly available Federal authority data.

### Data and code

The data and code underlying this manuscript are publicly available via zenodo https://doi.org/10.5281/zenodo.17508352 and GitHub https://github.com/KaMatthes/Spatial_temporal_morbidity_1918

## Acknowledgements

A special thanks goes to Kaspar Staub for previous/ongoing collaborations and helpful comments.

## Author contributions

Conceptualization: KM, SJ, RM

Data curation: KM, SJ,

Methodology: KM, SJ, RM

Data analysis: KM, SJ

Visualization: KM, SJ

Supervision: KM

Funding acquisition: KM

Writing – first draft: KM, SJ, RM

Writing – review & editing: KM, SJ, RM

## Supplementary material

**Supplementary Figure S1:**
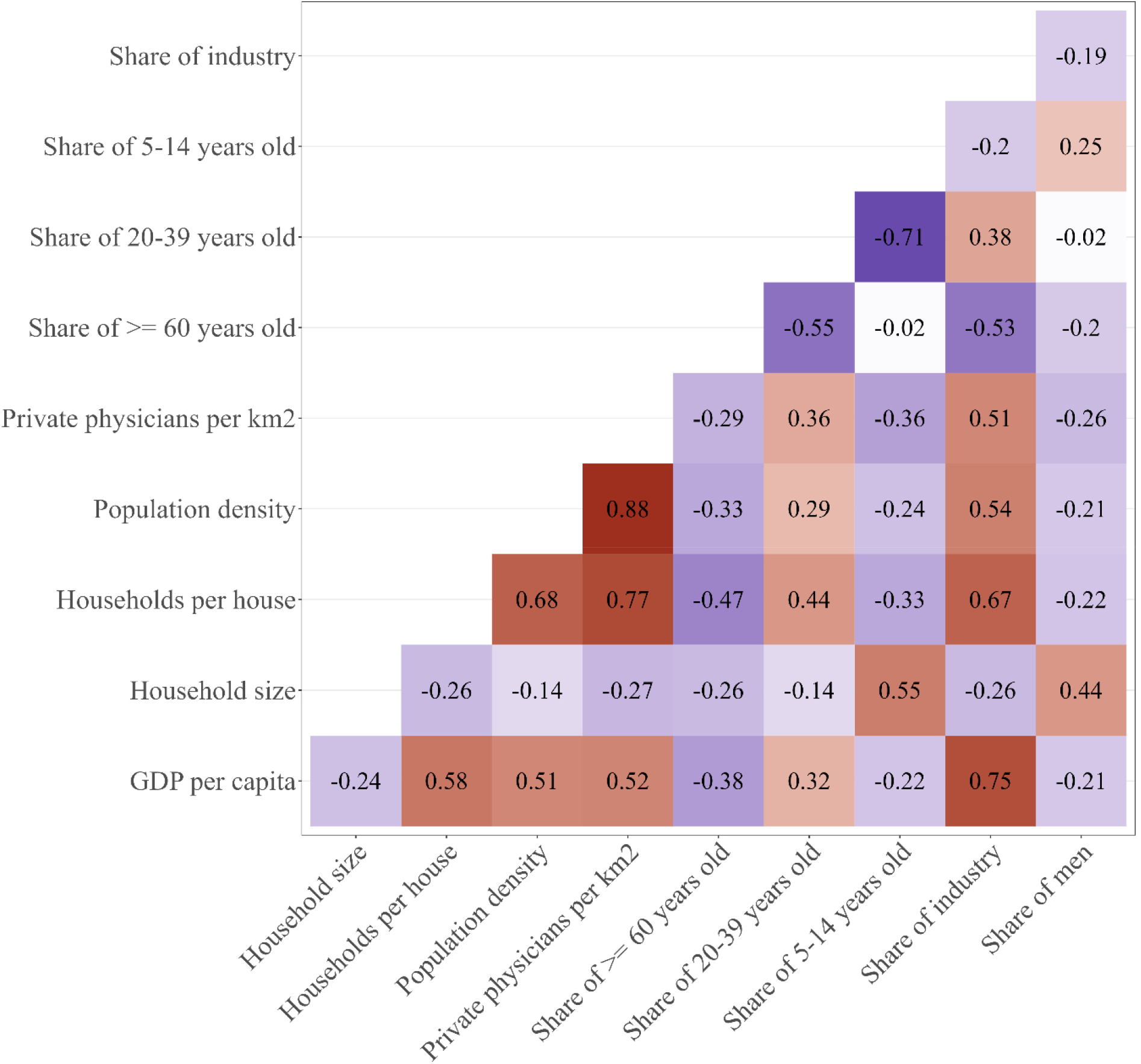
Correlation matrix of the explanatory variables

**Supplementary Figure S2:**
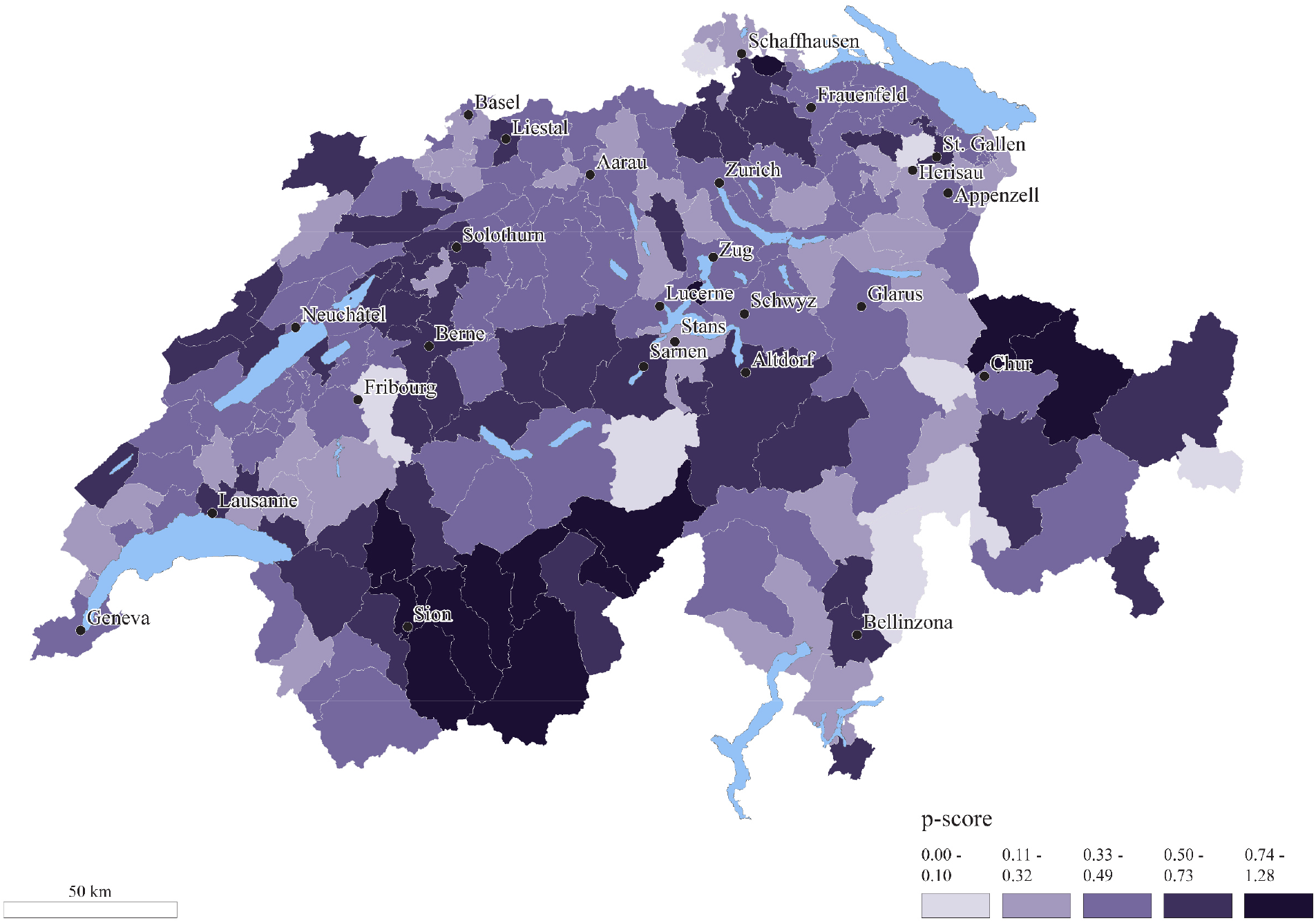
Excess mortality in each district.

**Supplementary Table S1:** Median regression coefficient of incidence and 95% confidence intervals (CI) from the robust GWR and a global linear regression. Each determinate is showing one model (univariable).

| Cofactor | Wave | GWR | Global |
| --- | --- | --- | --- |
| Population density | Jul 1918 - Aug 1918 | 21.22 (7.56 to 31.09) | 39.16 (12.97 to 65.34) |
| GDP per capita | Jul 1918 - Aug 1918 | 18.65 (11.36 to 27.97) | 50.44 (24.67 to 76.22) |
| Share of industry | Jul 1918 - Aug 1918 | 42.04 (31.36 to 50.06) | 66.23 (41.23 to 91.23) |
| Private physicians per km2 | Jul 1918 - Aug 1918 | 16.63 (2.46 to 22.64) | 24.94 (-1.61 to 51.5) |
| Households per house | Jul 1918 - Aug 1918 | 23.4 (14.15 to 33.95) | 28.12 (1.63 to 54.61) |
| Household size | Jul 1918 - Aug 1918 | 1.5 (-6.83 to 8.76) | 21.16 (-5.47 to 47.78) |
| Share of men | Jul 1918 - Aug 1918 | -14.64 (-20.38 to -4.98) | 11.84 (-14.91 to 38.58) |
| Share of 5-14 years old | Jul 1918 - Aug 1918 | 20.73 (11.56 to 26.98) | 47.63 (21.74 to 73.52) |
| Share of 20-39 years old | Jul 1918 - Aug 1918 | 25.42 (5.07 to 39.47) | -0.38 (-27.18 to 26.42) |
| Share of >= 60 years old | Jul 1918 - Aug 1918 | -34.78 (-41.52 to -20.08) | -51.8 (-77.52 to -26.08) |
| Population density | Sep 1918 - May 1919 | 10.74 (-42.48 to 34.56) | -22.89 (-151.39 to 105.62) |
| GDP per capita | Sep 1918 - May 1919 | 35.54 (3.1 to 53.23) | 15.64 (-112.89 to 144.16) |
| Share of industry | Sep 1918 - May 1919 | 41.27 (-3.31 to 83.05) | -25.94 (-154.44 to 102.55) |
| Private physicians per km2 | Sep 1918 - May 1919 | 22.17 (2.32 to 54.66) | 0.84 (-127.71 to 129.39) |
| Households per house | Sep 1918 - May 1919 | 57.03 (30.32 to 99.97) | -35 (-163.45 to 93.45) |
| Household size | Sep 1918 - May 1919 | -0.5 (-33.44 to 22.35) | -16.42 (-144.95 to 112.1) |
| Share of men | Sep 1918 - May 1919 | -24.39 (-100.88 to 19.32) | -84.74 (-212.69 to 43.22) |
| Share of 5-14 years old | Sep 1918 - May 1919 | -23.65 (-57.66 to 11.88) | -105.91 (-233.53 to 21.71) |
| Share of 20-39 years old | Sep 1918 - May 1919 | 34.63 (1.41 to 62.15) | 29.81 (-98.66 to 158.29) |
| Share of >= 60 years old | Sep 1918 - May 1919 | -28.28 (-57.16 to -4.51) | 120.33 (-7.02 to 247.67) |
| Population density | Jan 1920 - May 1920 | 40.74 (34.88 to 52.84) | 47.4 (15.71 to 79.1) |
| GDP per capita | Jan 1920 - May 1920 | 27.16 (18.33 to 39.59) | 38.53 (6.58 to 70.48) |
| Share of industry | Jan 1920 - May 1920 | 37.28 (23.09 to 48.82) | 40.94 (9.06 to 72.83) |
| Private physicians per km2 | Jan 1920 - May 1920 | 57.14 (42.58 to 71.99) | 29.96 (-2.18 to 62.11) |
| Households per house | Jan 1920 - May 1920 | 41.71 (32.16 to 56.48) | 35.38 (3.35 to 67.41) |
| Household size | Jan 1920 - May 1920 | -25.34 (-37.23 to -16.27) | -7.82 (-40.24 to 24.6) |
| Share of men | Jan 1920 - May 1920 | -38.26 (-42.96 to -34.09) | -37.45 (-69.43 to -5.47) |
| Share of 5-14 years old | Jan 1920 - May 1920 | -45.37 (-70.42 to -27.91) | -32.22 (-64.32 to -0.12) |
| Share of 20-39 years old | Jan 1920 - May 1920 | 71.05 (57.3 to 87.67) | 37.46 (5.48 to 69.44) |
| Share of >= 60 years old | Jan 1920 - May 1920 | -16.75 (-25.21 to -8.38) | -10.41 (-42.81 to 22) |

**Supplementary Table S2:**
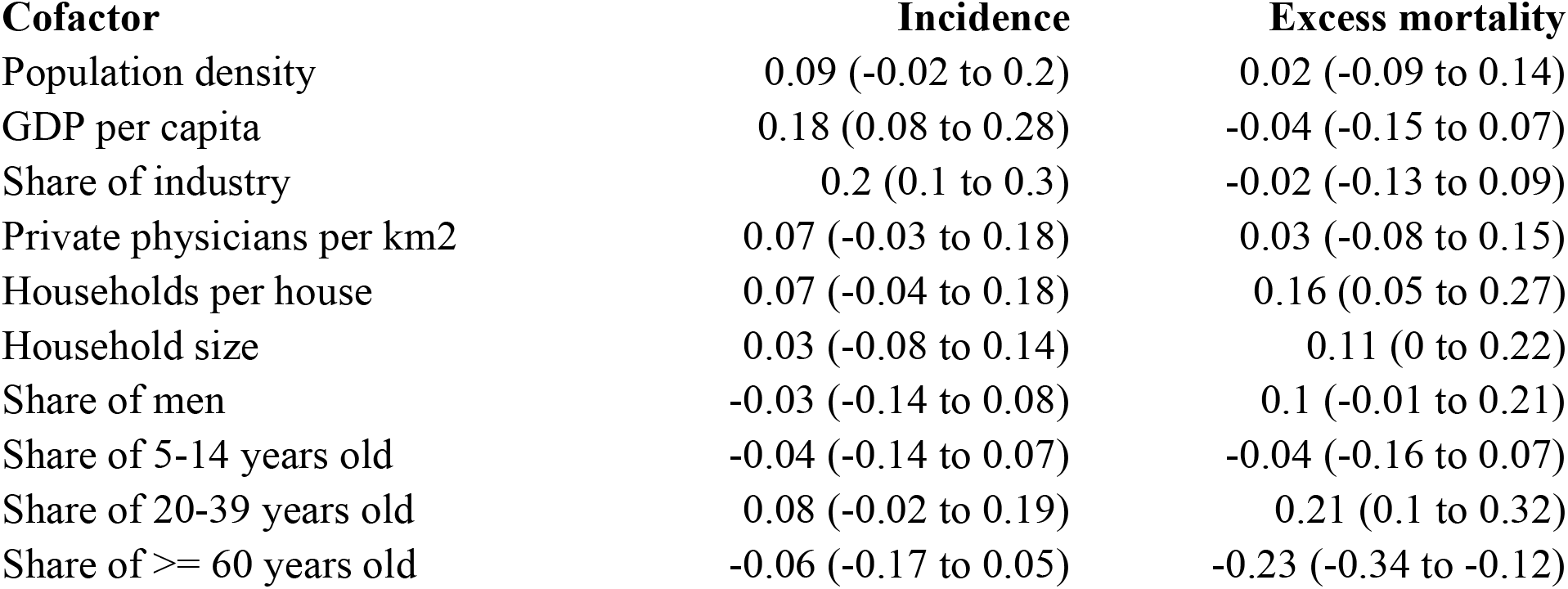
Regression coefficient and 95% confidence interval (CI) of the robust linear regression models. Each determinate is showing one model (univariable).

